# Distinct Autoimmune Antibody Signatures Between Hospitalized Acute COVID-19 Patients, SARS-CoV-2 Convalescent Individuals, and Unexposed Pre-Pandemic Controls

**DOI:** 10.1101/2021.01.21.21249176

**Authors:** Nahid Bhadelia, Anna C. Belkina, Alex Olson, Thomas Winters, Patricia Urick, Nina Lin, Ian Rifkin, Yachana Kataria, Rachel R. Yuen, Manish Sagar, Jennifer E. Snyder-Cappione

**Affiliations:** Department of Medicine, Boston University School of Medicine, Boston, MA, USA; National Emerging Infectious Diseases Laboratories (NEIDL), Boston University, Boston, MA, USA; Flow Cytometry Core Facility, Boston University School of Medicine, Boston, MA, USA; Department of Pathology and Laboratory Medicine, Boston University School of Medicine, Boston, MA, USA; Research Occupational Health Program, Boston University, Boston, MA, USA; Renal Section, Department of Medicine, Boston University School of Medicine, Boston, MA, USA; Renal Section, Department of Medicine, VA Boston Healthcare System, Boston, MA, USA; Department of Microbiology; Boston University School of Medicine, Boston, MA, USA

**Keywords:** SARS-CoV-2, COVID-19, autoantibodies, IgG, IgA, pre-pandemic, convalescent

## Abstract

Increasing evidence suggests that autoimmunity may play a role in the pathophysiology of SARS-CoV-2 infection during both the acute and ‘long COVID’ phases of disease. However, an assessment of autoimmune antibodies in convalescent SARS-CoV-2 patients has not yet been reported.

**Methodology:** We compared the levels of 18 different IgG autoantibodies (AABs) between four groups: (1) unexposed pre-pandemic subjects from the general population (n = 29); (2) individuals hospitalized with acute moderate-severe COVID-19 (n = 20); (3) convalescent SARS-COV-2-infected subjects with asymptomatic to mild viral symptoms during the acute phase with samples obtained between 1.8 and 7.3 months after infection (n = 9); and (4) unexposed pre-pandemic subjects with systemic lupus erythematous (SLE) (n = 6). Total IgG and IgA levels were also measured from subjects in groups 1-3 to assess non-specific pan-B cell activation.

**Results:** As expected, in multivariate analysis, AABs were detected at much higher odds in SLE subjects (5 of 6, 83%) compared to non-SLE pre-pandemic controls (11 of 29, 38%) [odds ratio (OR) 19.4,95% CI, 2.0 – 557.0, p = 0.03]. AAB detection (percentage of subjects with one or more autoantibodies) was higher in SARS-CoV-2 infected convalescent subjects (7 of 9, 78%) [OR 17.4, 95% CI, 2.0 – 287.4, p = 0.02] and subjects with acute COVID-19 (12 of 20, 60%) compared with non-SLE pre-pandemic controls, but was not statistically significant among the latter [OR 1.8,95% CI, 0.6 – 8.1, p = 0.23]. Within the convalescent subject group, AABs were detected in 5/5 with reported persistent symptoms and 2/4 without continued symptoms (p = 0.17). The multivariate computational algorithm Partial Least Squares Determinant Analysis (PLSDA) was used to determine if distinct AAB signatures distinguish subject groups 1-3. Of the 18 autoantibodies measured, anti-Beta 2-Glycoprotein, anti-Proteinase 3-ANCA, anti-Mi-2 and anti-PM/Scl-100 defined the convalescent group; anti-Proteinase 3-ANCA, anti-Mi-2, anti-Jo-1 and anti-RNP/SM defined acute COVID-19 subjects; and anti-Proteinase 3-ANCA, anti-Mi-2, anti-Jo-1, anti-Beta 2-Glycoprotein distinguished unexposed controls. The AABs defining SARS-COV-2 infected from pre-pandemic subjects are widely associated with myopathies, vasculitis, and antiphospholipid syndromes, conditions with some similarities to COVID-19. Compared to pre-pandemic non-SLE controls, subjects with acute COVID-19 had higher total IgG concentration (p-value=0.006) but convalescent subjects did not (p-value=0.08); no differences in total IgA levels were found between groups.

**Conclusions:** Our findings support existing studies suggesting induction of immune responses to self-epitopes during acute, severe COVID-19 with evidence of general B cell hyperactivation. Also, the preponderance of AAB positivity among convalescent individuals up to seven months after infection indicates potential initiation or proliferation, and then persistence of self-reactive immunity without severe initial disease. These results underscore the importance of further investigation of autoimmunity during SARS-CoV-2 infection and its role in the onset and persistence of post-acute sequelae of COVID-19.

## Introduction

The pathophysiology of acute COVID-19 appears to be a combination of damage due to direct viral invasion, coagulopathy, and an overactive and hyperinflammatory immune response (1-3). Evolving data is also now revealing that autoimmunity may be a critical component of the maladaptive immune reaction to SARS-CoV-2 infection (4). Recent studies show that SARS-CoV-2 infected individuals admitted to the hospital with more severe disease have higher rates of clinically relevant autoantibodies (AABs) (5, 6), with critically ill COVID-19 patients often possessing detectable levels of lupus anticoagulant antibodies, an antibody profile often implicated in hypercoagulable states (7). In addition, autoantibodies against type II interferons were found in COVID-19 subjects during acute illness (8); such antibodies may subvert the induction of an effective anti-SARS-CoV-2 immune response. Against the backdrop of this evolving autoimmune picture, an increasing number of reports have documented post-acute sequelae of COVID-19, or ‘long COVID’, found in those who survive acute illness yet continue to have prolonged symptoms ranging from sustained acute symptomatology, overall decrease in physiological reserve, and onset of new symptoms, including neurocognitive dysfunction (9, 10). In many cases, particularly those involving hospitalization and/or ventilation, the underlying pathophysiology of some of these symptoms could be the aftermath of severe acute illness itself and/or treatments; however, the prevalence of the ‘long COVID’ phenotype among individuals with initial mild disease implicates additional driving mechanisms (11, 12). We and others predict that events of acute infection, particularly immunological, may foreshadow the presence and pathophysiology of persistent symptoms (4). Currently, it remains unclear whether AABs found in acute COVID-19 subjects will remain post-convalescence, and if so, how long they persist and if such AABs track with long COVID phenotypes.

In this study, we examined the relative frequency and mean fluorescence intensity of 18 autoantibodies (AABs) from hospitalized patients with acute COVID-19, SARS-CoV-2 convalescent survivors up to 7 months after initial infection with varying symptom profiles, unexposed subjects from the general population collected before the pandemic, and pre-pandemic subjects with the autoimmune disease SLE. We performed multivariate computational analysis (Partial Least Squares Determinant Analysis (PLSDA)) to determine if distinct autoantibody signatures of the 18 measured distinguished COVID-19 subjects grouped by disease stage and severity with AABs found in the unexposed general population. Also, the total IgG and IgA levels were measured from most subjects as an indirect assessment of general B cell hyper-activation.

## Material and Methods

### Clinical Samples

Samples (plasma and serum) were collected and compared between four subject groups. First, retrospective pre-pandemic samples obtained from prior study cohorts which included both well controlled HIV seropositive and seronegative subjects from 2013-2019; this cohort is described in detail elsewhere (13). HIV+ subjects in this group (n=5) have undetectable viral loads for a minimum of 6 months with absolute CD4 T-cell counts above 300 cells/µL. Additionally, as a positive comparator group, banked study samples were obtained from lupus patients that fulfilled at least 4 of the 11 American College of Rheumatology revised criteria for the classification of SLE (14).

Banked acute COVID-19 samples and limited clinical dataset were obtained from a discarded clinical samples biorepository for patients admitted to the hospital (with or without ICU stay) during the spring and summer of 2020. These de-identified samples are from hospitalized patients at Boston Medical Center with confirmed PCR positivity for SARS-CoV-2.

SARS-CoV-2 infected convalescent survivors were prospectively recruited through anonymous blood collection study advertised through Boston University Research Occupational Health program. Survivors were identified and referred to the study by an occupational health physician who provided the patients with care during their initial illness and could confirm their initial COVID-19 PCR positivity. Participants in this study answered limited questions about demographics and dates and types of symptoms associated with COVID-19. Data regarding comorbidities was not available for this cohort.

### Ethics

Human Subjects review and permissions were obtained from the Boston University School of Medicine’s Institutional Review Board. Samples and minimal clinical data were deidentified at collection (COVID-19 survivors) or at sharing (pre-pandemic and acute COVID-19 cases).

### Autoantibody and total IgG and IgA detection

A multiplex assay was used to examine 18 AABs across the four study subgroups with MILLIPLEX MAP Human Autoimmune Autoantibody Panel kit (Cat. No. HAIAB-10K). We also measured total IgG with an IgG assay kit (ThermoFisher, Cat. BMS2091) and IgA levels were determined with an in-house ELISA protocol (‘BU ELISA’) previously described (15) with a recombinant human IgA1 used as the standard (Bio-Rad, Cat. HCA189) Total IgG and IgA measurements were performed for all acute and convalescent subjects as well as 11 randomly chosen non-SLE unexposed subjects. Quantification of analytes was performed on a Magpix (Luminex) instrument containing xPONENT 4.2 software (Boston University Analytical Core Facility) and a BioTek Synergy HT plate reader. Values reported were from analysis of almost all plasma samples except for serum obtained from two convalescent subjects. Matched analysis of plasma and serum from other convalescent subjects showed no differences in AAB values.

### Statistical Analysis

Among the pre-pandemic non-SLE control group, an average and standard deviation (STD) was calculated for the mean fluorescence intensity (MFI) for all 18 antibodies individually. An individual was classified as having an AAB if their MFI for a particular AAB was at least 3 STD above the pre-pandemic average. Frequencies of AABs among the groups were compared using Chi square. Multivariate logistic and Poisson linear regression was used to determine the odds of possessing an AAB and the number of AABs respectively. In these multivariate analyses, predictors included the different groups (SLE, convalescent and acute COVID patients relative to the control individuals), age, gender, HIV status, and black race versus other race/ethnicity. Partial Least Squares discriminant analysis (PLSDA) was conducted using PLS Toolbox (Eigenvector Research, Inc.) package for MATLAB (MathWorks) to test whether a set of autoantibodies can distinguish between three clinical groups (acute, convalescent and non-SLE unexposed). Raw fluorescence data was normalized by Z-score before application of the algorithm. The importance of each parameter to the overall model prediction was quantified using variable importance in projection (VIP) score. To eliminate noise in the data, the first pass analysis calculated VIP scores for 18 measured autoantibodies (complete dataset) and allowed to enrich for 10 autoantibodies with VIP scores > 0.9 in a preliminary model. Then, a PLSDA model was built based on these 10 autoantibodies. Cross-validation was performed with one-third of the dataset and final VIP scores were calculated. The number of latent variables (LVs) was chosen so as to minimize cumulative error over all predictions. A VIP score >1 (above average contribution) was considered important for model performance and prediction. Model confidence was calculated by randomly permuting Y 100 times and rebuilding the model to form a distribution of error for these randomly generated models, then comparing the model to this distribution with the Mann–Whitney U-test to determine the significance of the model. Statistical analyses were performed using GraphPad version 9 or Matlab. All p-values are 2-sided unless indicated.

## Results

To examine possible connections between SARS-CoV-2 infection the presence and signature of autoimmune antibodies (AABs), plasma and/or serum was collected from four subject groups: (1) unexposed subjects with samples collected prior to fall 2019 (n=29), with an average age was 55.7 years, and 21 (72.4%) were male; (2) acutely ill hospitalized patients between April and June 2020 with moderate or severe COVID-19 (n=20), with an average age of 62.6 years, and 16 (80%) were male, with samples were collected 3-40 days after onset of symptoms, and 12 (60%) individuals in this cohort required intensive care unit level care during their admission, one was HIV+, and no patients died; (3) SARS-COV-2 infected convalescent subjects (n=9), enrolled between 1.8 and 7.3 months after their initial viral diagnosis, with none were hospitalized during the acute course of their infection,. with an average age was 53 years, and two (22%) were male. Persistent or prolonged symptoms in the convalescent group was defined as any symptoms lasting more than one month after diagnosis; this was reported by 55% (5/9) of the individuals and one individual remained asymptomatic (subject S2, Figure 1A and Supplemental Table 1). Those convalescent subjects with prolonged symptoms reported either persistence of acute symptoms such as cough, loss of sense of smell or taste, shortness of breath, or onset of new symptoms such as ringing of the ears. The final subject group (4) was comprised of individuals with Systemic Lupus Erythematosus (SLE) subjects (n=6); all samples were collected prior to the pandemic, with an average age of 46.5, of which 3 (50%) were male (Table 1).

**Table 1:** Description of Study Groups.

| **Table 1: Description of Study Groups** |  |  |  |  |
| --- | --- | --- | --- | --- |
|  | **Pre-pandemic** | **Acute COVID-19** | **COVID-19 Survivors** | **SLE** |
|  | (n/%) | (n/%) | (n/%) | (n/%) |
| N (%)= 64 | 29 (45.3) | 20 (31.2) | 9(14) | 6(9.4) |
| Age (mean) [years] | 55.7 | 62.6 | 53 | 46.5 |
| Sex (n/%) [male] | 21 (72.4) | 16 (80) | 2 (20) | 3(50) |
| Race/Ethnicity (n/%) |  |  |  |  |
| *White* | 13 (44.8) | 2(10) | 6(66.7) | 2(33.3) |
| *Black* | 16 (55.2) | 11(55) | 3(33.7) | 1(16.6) |
| *Hispanic* | 0 | 1(5) | 0 | 1(16.6) |
| *Asian* | 0 | 1(5) | 0 | 1(16.6) |
| *Unknown/other* | 0 | 4(20) | 0 |  |
| HIV seropositive (n/%) | 5(17.2) | 1(5) | n/a | n/a |
| ICU admission | n/a | 12(60) | n/a | n/a |
| Days since COVID19 symptom onset (n/%) | n/a |  |  | n/a |
| ≤10 days |  | 13(65) |  |  |
| ≤30 days |  | 6(30) |  |  |
| ≤60 days |  | 1(5) | 1(11.1) |  |
| ≤3 months |  |  | 2(22.2) |  |
| ≤6 months |  |  | 1(11.1) |  |
| >6months |  |  | 5(55.6) |  |
| Presence of persistent symptoms | n/a | n/a | 5(55.6) | n/a |
| *Date of diagnosis was used for one asymptomatic COVID-19 survivor |  |  |  |  |

**Figure 1.**
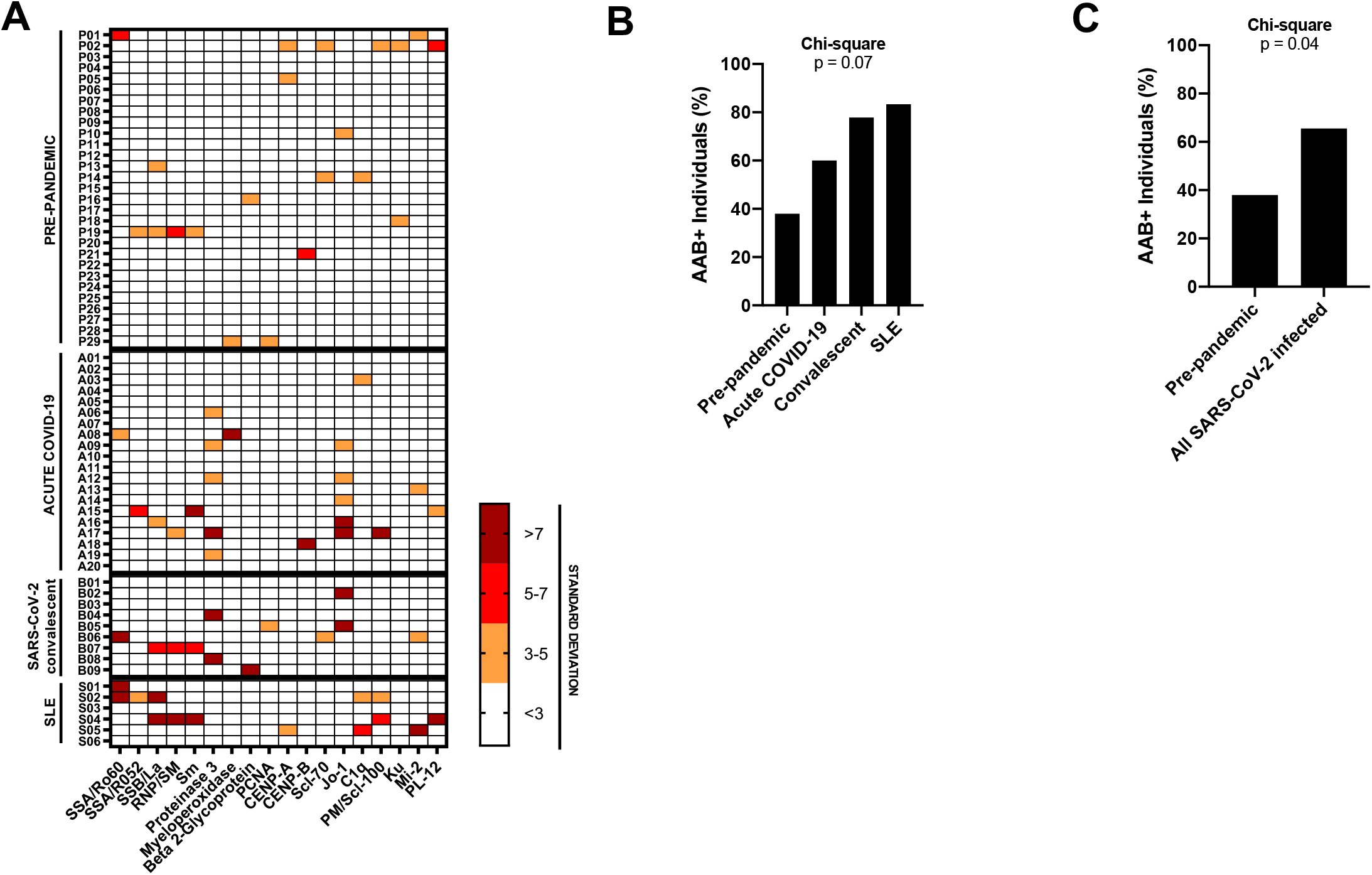
Autoantibody (AAB) expression among SARS-CoV-2 acute, convalescent groups compared with pre-pandemic non-autoimmune controls. (A) Heat map sorted by study groups indicate positive autoantibody hits compared to mean of Raw MFI values from pre-pandemic controls. Positive hits were considered greater than 3 standard deviations (SD) from mean as indicated by orange (3-5 SD), red (5-7 SD), and dark red (>7 SD) squares. White squares represent values less than 3 SD. Raw MFI values available in supplementary Table 1. (B) The percentage of subjects with at least one positive autoantibody hit is shown for all four subject groups (Chi Square, p = 0.07), and pre-pandemic and all SARS-COV-2 infected (acute COVID-19 and convalescent) subjects with at lest one AAB is compared (C). Autoantibody abbreviations: SSA/Ro60= Anti-Sjögren’s Syndrome-related antigen A/Ro60 kDa; SSA/Ro52=Anti-Sjögren’s Syndrome-related antigen A/Ro52 kDa; SSB/La= Anti-Sjögren’s Syndrome-related antigen B/La; RNP/Sm= Anti-RNP/Smith; Sm= anti-Sm; Prot 3= Anti-Proteinase 3; Myeloper= Anti-Myeloperoxidase; Beta 2-G= Anti-β 2-Glycoprotein; CENP-A= Anti-Centromere Protein A; CENP-B= Anti-Centromere Protein B; Scl-70= Anti-Scl-70; Jo-1= Anti-Jo-1; C1q= Anti-C1q;PM/Scl-100=Anti-PM/Scl-100; Ku= Anti-Ku; Mi2= Anti-Mi-2; PL-12= Anti-Alanyl-tRNA synthetase

The mean fluorescence intensity (MFI) of 18 autoantibodies (AABs) were measured from all subject samples, with positive hits defined as a MFI value greater than the average +3SD of all pre-pandemic samples for each AAB readout (Figure 1A, Supplemental Table 1). At least one AAB was present in 55% (11/20) of acute COVID-19 subjects and 78% (7/9) of SARS-COV-2 infected convalescent survivors samples, both groups showing higher prevalence than pre-pandemic group 1 (38%, 11/29); 83% (5/6) of SLE samples were AAB+ (Chi-square p-value across all groups=0.07) (Figures 1A, B). AAB positivity among all 29 SARS-CoV-2 infected subjects was 66%, nearly twice the level of the pre-pandemic group 1 (Figure 1C) (Chi-square p-value=0.04). In multivariate analysis, the odds ratios for detected AABs were 19.4 (95% CI, 2.0 – 557.0, p-value 0.03) for SLE samples, 1.8 (95% CI, 0.6 – 8.1, p-value=0.23) for acute COVID-19 samples and 17.4 (95% CI, 2.0 – 287.4, p-value=0.02) for convalescent samples, relative to the pre-pandemic group 1. Poisson linear regression showed that SLE and convalescent but not the acute COVID group were associated with significantly higher number of AABs relative to the pre-pandemic non-SLE controls (Table 2).

**Table 2:** Multivariate Analysis of Correlation of Total Amount of Autoimmune Antibodies Compared to Non-SLE Pre-pandemic Samples.

|  |  |  |  |
| --- | --- | --- | --- |
| Study Group | $\beta$ | 95% CI | p-value |
| SLE | 2.0 | 1.0 – 3.0 | <0.0001 |
| Acute Infection | 0.57 | 0.10 – 1.3 | 0.1 |
| Survivor | 1.16 | 0.25 – 2.1 | 0.01 |

Interestingly, AABs were detected in all 5 convalescent individuals who reported persistent symptoms (S4, S6, S7, S8, S9) In contrast, AABs were only detected in 2 of the 4 convalescent individuals without symptoms beyond acute phase of illness (S1, S2, S3, S5) (p value=0.17). Among the acute subjects, individuals who required intensive care (8/12) were not more likely to have AABs detected compared to those who did not (4/8), (p value=0.65).

We next used PLSDA (partial least squares determinant analysis) to test whether a subset of autoantibodies can distinguish between the pre-pandemic non-SLE (group 1), acute COVID-19 (group 2) and SARS-CoV-2 infected convalescent (group 3) subjects. PLSDA determines which measurements from each subject would “fit” that subject into its clinical cohort and verifies if such classification is statistically significant. We sequestered a subset of 10 AABs that generated >0.9 VIP (variable importance in projection, see Methods) scores in the model, suggesting they may contribute to separation of three subject groups of interest (Figure 2). These autoantibodies were incorporated into a PLSDA model whose confidence per permutation test was 81% for survivor group, 89% for acute group, and 99% for pre-pandemic group with 66% CV accuracy (i.e. two-fold better than random distribution). We identified autoantibodies that contribute to separation of the groups per their VIP scores, where scores over 1 indicate that the variable significantly contributes to separation. The AABs that separated patients with acute COVID-19 were anti-RNP/anti-Smith, anti-proteinase 3-ANCA, Anti-Jo-1, Ant-Mi-2, whereas those that separated survivors were Proteinase 3, Beta 2-Glycoprotein, PM/Scl-100 and MI-2. The MFI of these AABs was also higher in the acute and convalescent groups compared to pre-pandemic non-SLE controls as previously mentioned (Supplemental Table 1).

**Figure 2.**
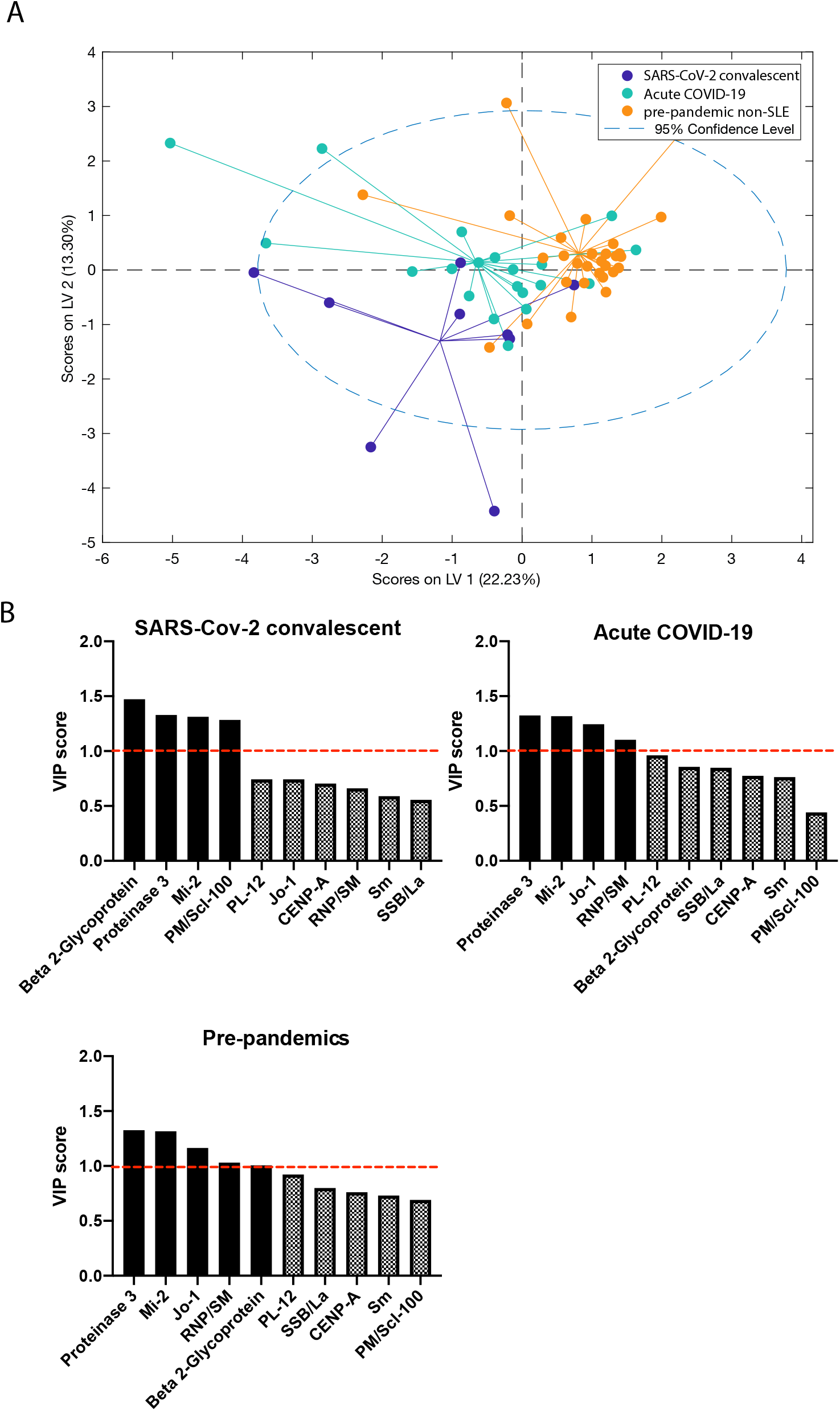
Partial Least Squares Discriminant Analysis (PLSDA) of autoimmune antibodies from Acute COVID-19, convalescent SARS-CoV-2 infected, and pre-pandemic controls. (A) PLSDA model distinguished autoantibody profiles for Acute COVID 19 (teal), convalescent SARS-CoV-2 infected (blue) and pre-pandemic (orange) subjects with 66% cross-validated prediction accuracy. B, C, D Variable importance in projection (VIP) scores of autoantibodies defining each cohort. Autoantibodies with above average contribution to differences between cohort profiles (VIP >1) are shown as solid bars.

To assess if the higher percentages of subjects with autoantibodies in SARS-CoV-2 infected groups compared with pre-pandemics (group 1) is a reflection of general B cell hyper-activation, total IgG and IgA levels were measured. Pre-pandemic group 1 and SARS-CoV-2 convalescent groups (p value =0.08) possessed lower circulating IgG compared with the acute COVID-19 subjects, with the difference between pre-pandemic and acute COVID-19 showing statistical significance (p value=0.0063) (Figure 3A); IgA levels were similar between the groups (p value=0.09 and p value=0.26) (Figure 3B).

**Figure 3A and 3B.**
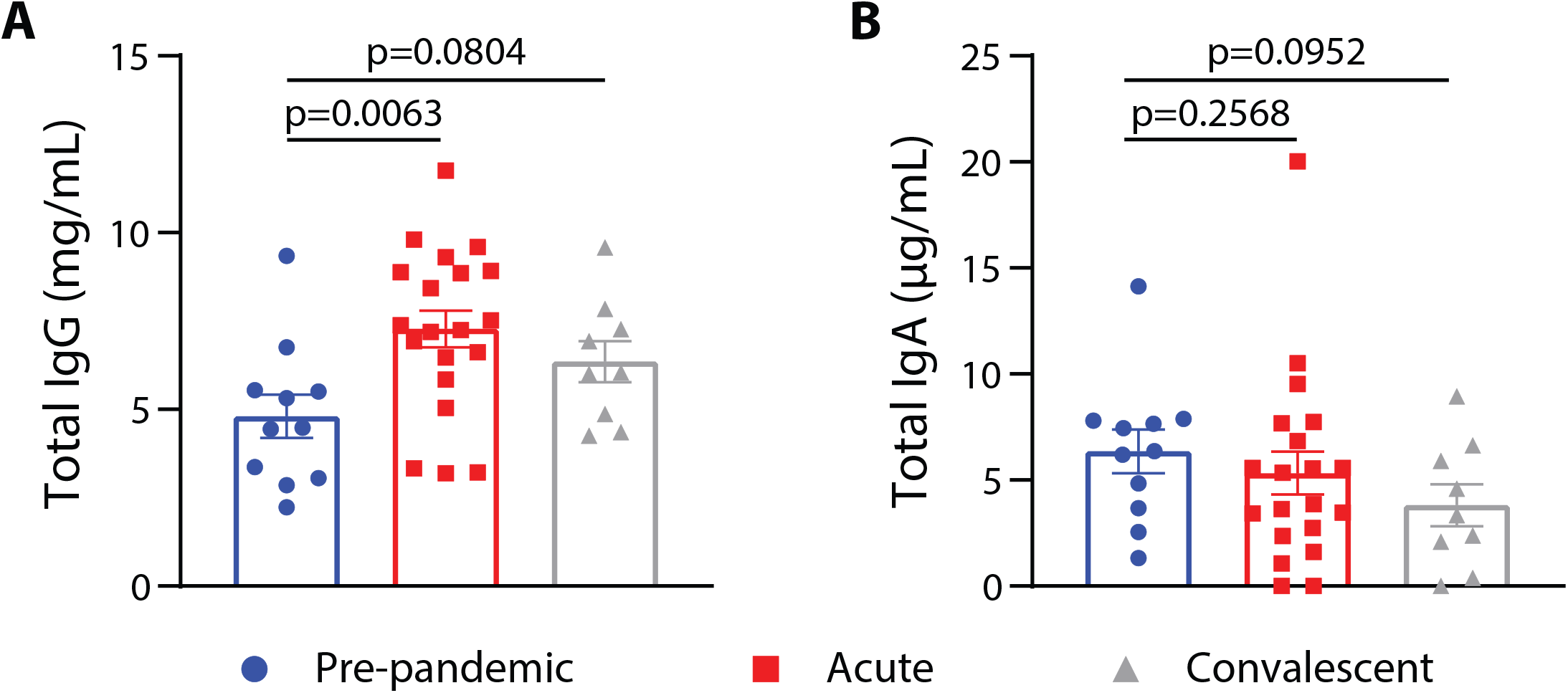
Total circulating IgG and IgA concentrations from pre-pandemic non-SLE (group 1), acute hospitalized COVID-19 (group 2), and SARS-COV-2 convalescent (group 3)subjects. Values for each subject in each group are shown as symbols. Unpaired non-parametric Mann-Whitney t-tests were performed to determine significance between the groups.

Values reported were from analysis of almost all plasma samples except for serum obtained from two convalescent subjects. Matched analysis of plasma and serum from other convalescent subjects showed no differences in AAB values.

## Discussion

Here, we report a majority of SARS-COV-2 convalescent individuals with asymptomatic-mild acute disease possess at least one autoimmune antibody (AAB) (7/9, 83%). Our results also confirm findings from other studies that have demonstrated high levels of autoimmune antibodies (AABs) in those with acute COVID-19, and we demonstrate that these levels are again higher than what was noted in pre-pandemic controls representing the general population but not statistically significant (5, 6).

We did not find statically significant correlation between disease severity among hospitalized acute subjects (need for intensive level care) and detection of AABs but our study was not powered to control for other recognized risk factors for COVID-19 disease severity (16). Our study was also not designed or powered to correlate presence of persistent symptoms in convalescent phase with detection of AABs but finding of such antibodies in all subjects with persistent symptoms highlights importance of further study in a larger cohort.

If antibodies against SARS-CoV-2 generated during acute illness cross react with human (self) epitopes, it suggests that they persist for a period of time after recovery and may contribute to symptoms noted in convalescence. However, whether such increases in AABs are pathogenic and related to persistent symptoms in patients with SARS-CoV-2 or whether they represent part of overall B cell proliferation in the aftermath of the infection has remained unclear.

We demonstrated that patients with acute COVID-19 had elevated immunoglobulin (IgG but not IgA) which may be a reflection of polyconal B cell activation in response to sepsis-like conditions, in agreement with a prior study (17). It remains unclear if increased AABs detected in acute are a reflection of this overall chronic, nonspecific activation. A similar increase in IgG was not noted in convalescent samples despite higher odds detecting AABs in this group.

Low titers of circulating titer AABs, including antiphospholipid antibodies, generated in the aftermath of viral infections have been documented with numerous pathogens united Epstein-Barr virus, Parvovirus B19 and Cytomegalovirus among other (18-21). In most patients, these titers tend to decline over time and the role of post viral infection induced AABs in any downstream pathological manifestations have remained unclear (22-24). Like other viral infections, acute illness with COVID-19 may induce autoimmunity through molecular mimicry, ‘bystander activation’ during a hyperinflammatory state, or epitope spreading and presentation of cryptic antigens due to excessive cell damage. Alternatively, there may be a rapid autoimmune or autoinflammatory dysregulation, based on genetic predisposition, which would explain why some patients mount a detrimental initial response (25).

Although AABs are detected at the population level in disease-free individuals at varying prevalence rates, a recent analysis showed that hexapeptides from immunoreactive epitopes present in SARS-CoV-2 are widespread and shared among a high number of human proteins, spanning numerous organ systems (26-29). Similarly, the AABs identified as being most closely correlated with and elevated in the acute and convalescent samples in our cohort are varied and have previously been associated with myopathies, dermatomyosities, vasculitides and anti-phospholipid syndromes (30-33).

There are several limitations to our study including small sample size and the lack of longitudinal data from individual COVID-19 patients. The latter did not allow internal control to examine the dynamics of the autoimmune response over time or to control for preexistence of subclinical circulating AABs among participants before COVID-19 infection. Additionally, we had limited data on comorbidities for COVID-19 survivors and could not control for whether they had existing diagnosis of an autoimmune disorder. We also had limited data on concurrent medical comorbidities among subjects in our pre-pandemic sample. In our very small cohort of COVID-19 survivors, over half of the participants reported persistent or prolonged symptoms after acute infection lasting over one month. Bias may also have been introduced by who responded to recruitment for the anonymous blood collection study, if those suffering with persistence symptoms were more likely to agree to participate. We had only serum samples for two COVID-19 survivor participants but by performing matched analysis on other samples, we were able to demonstrate that this did not affect the measurement of AABs we observed.

Despite these limitations, the discovery of AABs at such high levels in even a small, sampled group of convalescent subjects compared to pre-pandemic controls in this study is a significant finding worthy of further evaluation. Future investigations should attempt to correlate whether “long COVID” features with presence of autoimmune antibodies to identify whether autoimmunity places a role in persistent symptoms or prolonged recovery. If on the other hand, subclinical circulating AABs represent a propensity for greater autoimmune response during acute infection or serve as a marker for more severe disease, it would be critical correlate preexistence of and discern types of AABs associated with detrimental clinical course during acute infections and after recovery. Patients with systemic autoimmune diseases do not, at least at this time, seem to be at higher risk for acquiring SARS-CoV-2 infections (34). Lastly, further studies should correlate the dynamics over time of AABs in patients with SARS-CoV-2 infections compared to pathogen specific antibodies created during acute illness.

## Conclusion

We demonstrated that autoimmune antibodies are present in convalescent subjects with prior mild SARS-CoV-2 infection up to over seven months into convalescence. The significance of these findings remain unclear as does their correlation to the clinical phenotype of “long COVID.” Additional research from larger cohort needs to focus on autoimmunity as one possible aspect of pathophysiology leading to persistent physiological symptoms noted in many COVID-19 survivors. Additionally, our findings support other studies reporting the presence of immune responses to self-epitopes during acute SARS-CoV-2 infection. New investigations must elucidate if these epitopes are revealed during illness or whether propensity or presence of circulating autoimmune antibodies place some patients at higher risk for worse COVID-19 outcomes.

## Data Availability

The authors confirm that the data supporting the findings of this study are available within the article [and/or] its supplementary materials.

