## Supplemental Table 1 for "Distinct Autoimmune Antibody Signatures Between Hospitalized Acute COVID-19 Patients, SARS-CoV-2 Convalescent Individuals, and Unexposed Pre-Pandemic Controls"

**Supplemental Table I. Autoantibody profiles from unexposed pre-pandemic, acute COVID-19 hospitalized, SARS-CoV-2 infected convalescent, and unexposed SLE subjects.** The Mean Fluorescence Intensity (MFI) of each result is shown. All colored boxes indicate the presence of AAB with positivity and magnitude determined individually for each readout as follows : yellow = MFI > the average of all non-SLE pre-pandemic subjects (group 1, n=29) +3 standard deviations (SD) of that value, orange = MFI > avg of non-SLE pre-pandemics +5 SD, red => avg of non-SLE pre-pandemics +8 SD. Individual subejcts with at least one AAB have labels in bold. Autoantibody abbreviations: SSA/Ro60= Anti-Sjögren's Syndrome-related antigen A/Ro60 kDa; SSA/Ro52=Anti-Sjögren's Syndrome-related antigen A/Ro52 kDa; SSB/La= Anti-Sjögren's Syndrome-related antigen B/La; RNP/Sm= Anti-RNP/Smith; Sm= anti-Sm; Prot 3= Anti-Proteinase 3; Myeloper= Anti-Myeloperoxidase; Beta 2-G= Anti-β 2-Glycoprotein; CENP-A= Anti-Centromere Protein A; CENP-B= Anti-Centromere Protein B; Scl-70= Anti-Scl-70; Jo-1= Anti-Jo-1; C1q= Anti-C1q; PM/Scl-100=Anti-PM/Scl-100; Ku= Anti-Ku; Mi2= Anti-Mi-2; PL-12= Anti-Alanyl-tRNA synthetase

| | Autoantibody | | | | | | | | | | | | | | | | | |
|---|---|---|---|---|---|---|---|---|---|---|---|---|---|---|---|---|---|---|
| | SSA/Ro60 | SSA/R052 | SSB/La | RNP/SM | Sm | Prot 3 | Myeloper | Beta 2-G | PCNA | CENP-A | CENP-B | Scl-70 | Jo-1 | C1q | PM/Scl-100 | Ku | Mi-2 | PL-12 |
| **Pre-pandemic** | | | | | | | | | | | | | | | | | | |
| **P1** | 1394 | 1116 | 15 | 5 | 27 | 45 | 8 | 22 | 1031 | 113 | 7 | 233 | 271 | 57 | 184 | 209 | 1120 | 182 |
| **P2** | 41 | 4110 | 81 | 45 | 241 | 58 | 28 | 52 | 213 | 20183 | 1911 | 6858 | 278 | 882 | 1010 | 10300 | 643 | 3632 |
| P3 | 24 | 245 | 18 | 6 | 109 | 193 | 166 | 40 | 17 | 313 | 1280 | 73 | 33 | 17 | 11 | 159 | 88 | 49 |
| P4 | 81 | 111 | 23 | 9 | 78 | 61 | 21 | 66 | 104 | 202 | 92 | 147 | 381 | 28 | 185 | 1081 | 445 | 176 |
| **P5** | 36 | 8103 | 50 | 27 | 182 | 201 | 21 | 23 | 35 | 19519 | 836 | 522 | 297 | 347 | 68 | 875 | 162 | 146 |
| P6 | 36 | 217 | 76 | 26 | 356 | 57 | 19 | 85 | 23 | 284 | 104 | 104 | 59 | 24 | 58 | 121 | 200 | 122 |
| P7 | 36 | 709 | 18 | 5 | 54 | 35 | 5 | 356 | 17 | 45 | 11 | 163 | 35 | 1549 | 10 | 84 | 69 | 196 |
| P8 | 17 | 115 | 6 | 51 | 50 | 24 | 8 | 52 | 19 | 143 | 2 | 301 | 64 | 45 | 195 | 130 | 101 | 76 |
| P9 | 21 | 80 | 20 | 8 | 84 | 24 | 36 | 26 | 555 | 556 | 189 | 109 | 785 | 96 | 18 | 2630 | 82 | 59 |
| **P10** | 21 | 163 | 60 | 32 | 263 | 29 | 13 | 97 | 3160 | 126 | 11 | 55 | 3195 | 173 | 22 | 36 | 83 | 62 |
| P11 | 47 | 493 | 24 | 17 | 269 | 35 | 8 | 8 | 388 | 44 | 10 | 426 | 69 | 5570 | 744 | 28 | 386 | 45 |
| P12 | 40 | 85 | 28 | 17 | 81 | 159 | 10 | 45 | 35 | 160 | 27 | 322 | 1018 | 342 | 137 | 220 | 139 | 247 |
| **P13** | 31 | 88 | 584 | 10 | 59 | 35 | 3 | 12 | 934 | 53 | 4 | 94 | 29 | 107 | 58 | 164 | 80 | 66 |
| **P14** | 204 | 349 | 223 | 80 | 531 | 85 | 23 | 87 | 130 | 1704 | 648 | 6536 | 366 | 13272 | 49 | 229 | 222 | 304 |
| P15 | 8 | 45 | 3 | 4 | 15 | 17 | 3 | 283 | 17 | 787 | 29 | 42 | 686 | 459 | 10 | 119 | 74 | 39 |
| **P16** | 11 | 54 | 108 | 8 | 29 | 28 | 9 | 1003 | 2763 | 686 | 272 | 84 | 29 | 57 | 27 | 52 | 270 | 55 |
| P17 | 15 | 1622 | 215 | 11 | 29 | 62 | 17 | 69 | 28 | 3725 | 22 | 52 | 71 | 49 | 104 | 44 | 236 | 126 |
| **P18** | 37 | 786 | 39 | 25 | 103 | 43 | 15 | 152 | 26 | 18091 | 26 | 80 | 69 | 327 | 829 | 13335 | 155 | 390 |
| **P19** | 35 | 20409 | 689 | 465 | 1608 | 104 | 26 | 127 | 1396 | 702 | 31 | 220 | 437 | 693 | 318 | 1913 | 150 | 663 |
| P20 | 43 | 73 | 26 | 18 | 72 | 53 | 18 | 28 | 27 | 94 | 10 | 1452 | 29 | 163 | 116 | 73 | 108 | 470 |
| **P21** | 18 | 149 | 40 | 17 | 86 | 183 | 74 | 65 | 30 | 1064 | 12100 | 127 | 529 | 2623 | 31 | 414 | 615 | 124 |
| P22 | 106 | 382 | 116 | 68 | 538 | 86 | 19 | 83 | 109 | 451 | 27 | 250 | 217 | 3974 | 603 | 751 | 389 | 1185 |
| P23 | 16 | 64 | 47 | 18 | 225 | 32 | 7 | 16 | 8 | 167 | 8 | 94 | 37 | 21 | 21 | 21 | 58 | 37 |
| P24 | 9 | 408 | 71 | 10 | 76 | 197 | 11 | 21 | 17 | 139 | 10 | 48 | 53 | 590 | 19 | 357 | 63 | 22 |
| P25 | 73 | 84 | 22 | 15 | 55 | 155 | 18 | 40 | 28 | 93 | 48 | 3256 | 168 | 566 | 87 | 579 | 75 | 44 |
| P26 | 17 | 47 | 36 | 18 | 33 | 82 | 48 | 36 | 130 | 143 | 21 | 42 | 236 | 40 | 52 | 119 | 54 | 318 |
| P27 | 24 | 332 | 11 | 6 | 57 | 54 | 3 | 758 | 19 | 397 | 23 | 37 | 223 | 220 | 17 | 51 | 63 | 56 |
| P28 | 10 | 134 | 31 | 12 | 60 | 41 | 3 | 12 | 621 | 915 | 10 | 90 | 215 | 25 | 18 | 205 | 87 | 68 |
| **P29** | 12 | 336 | 20 | 7 | 29 | 194 | 225 | 85 | 7018 | 83 | 7 | 39 | 186 | 85 | 182 | 4881 | 66 | 118 |
| **Acute COVID-19, hospitailzed** | | | | | | | | | | | | | | | | | | |
| A1 | 5 | 41 | 5 | 6 | 17 | 25 | 1 | 14 | 39 | 687 | 2 | 22 | 43 | 10 | 65 | 48 | 51 | 21 |
| A2 | 69 | 1272 | 339 | 147 | 643 | 204 | 20 | 110 | 548 | 755 | 40 | 167 | 1058 | 66 | 51 | 336 | 315 | 828 |
| **A3** | 58 | 112 | 65 | 61 | 144 | 217 | 40 | 179 | 123 | 164 | 131 | 1796 | 470 | 13944 | 90 | 1062 | 233 | 389 |
| A4 | 12 | 94 | 488 | 11 | 136 | 35 | 7 | 51 | 10 | 8601 | 253 | 947 | 134 | 24 | 11 | 42 | 33 | 33 |
| A5 | 49 | 149 | 91 | 54 | 223 | 139 | 19 | 46 | 480 | 674 | 619 | 404 | 176 | 89 | 37 | 99 | 551 | 140 |
| **A6** | 35 | 308 | 89 | 52 | 333 | 280 | 151 | 121 | 4150 | 890 | 26 | 309 | 541 | 1206 | 37 | 212 | 279 | 496 |
| A7 | 37 | 1264 | 333 | 189 | 811 | 183 | 14 | 169 | 234 | 267 | 293 | 111 | 270 | 100 | 40 | 2152 | 97 | 252 |
| **A8** | 886 | 498 | 269 | 171 | 885 | 123 | 3349 | 454 | 16 | 184 | 24 | 1042 | 171 | 107 | 24 | 32 | 91 | 96 |
| **A9** | 101 | 2474 | 76 | 49 | 139 | 291 | 16 | 31 | 113 | 629 | 73 | 763 | 3179 | 120 | 911 | 4092 | 535 | 662 |
| A10 | 21 | 3178 | 33 | 19 | 66 | 89 | 10 | 457 | 32 | 233 | 40 | 222 | 326 | 546 | 36 | 130 | 121 | 187 |
| A11 | 28 | 885 | 66 | 46 | 163 | 157 | 31 | 68 | 47 | 270 | 22 | 140 | 1209 | 194 | 114 | 990 | 688 | 234 |
| **A12** | 58 | 498 | 228 | 142 | 516 | 263 | 56 | 77 | 30 | 315 | 386 | 353 | 2221 | 107 | 75 | 61 | 122 | 477 |
| **A13** | 247 | 279 | 45 | 26 | 52 | 99 | 20 | 203 | 87 | 2008 | 24 | 954 | 105 | 172 | 41 | 214 | 893 | 127 |
| **A14** | 27 | 624 | 100 | 43 | 131 | 70 | 11 | 16 | 193 | 1178 | 20 | 2344 | 2064 | 343 | 52 | 817 | 272 | 397 |
| **A15** | 35 | 21600 | 48 | 110 | 8031 | 68 | 162 | 120 | 245 | 357 | 82 | 770 | 1197 | 2203 | 206 | 81 | 246 | 2811 |
| **A16** | 149 | 979 | 722 | 120 | 522 | 229 | 61 | 220 | 158 | 2240 | 703 | 891 | 10365 | 869 | 179 | 2844 | 388 | 1286 |
| **A17** | 149 | 380 | 246 | 326 | 199 | 656 | 68 | 542 | 239 | 1376 | 27 | 3117 | 11806 | 146 | 1997 | 3874 | 451 | 1930 |
| **A18** | 507 | 218 | 109 | 87 | 142 | 191 | 56 | 233 | 4126 | 223 | 31373 | 423 | 126 | 400 | 112 | 1617 | 326 | 628 |
| **A19** | 55 | 574 | 148 | 104 | 676 | 278 | 48 | 214 | 63 | 2214 | 44 | 1633 | 1271 | 931 | 175 | 9230 | 508 | 2033 |
| A20 | 46 | 341 | 81 | 65 | 106 | 230 | 85 | 512 | 2178 | 298 | 453 | 455 | 420 | 157 | 94 | 1021 | 390 | 1651 |
| **SARS-CoV-2 Convalescent** | | | | | | | | | | | | | | | | | | |
| S1 | 22 | 53 | 28 | 10 | 65 | 99 | 15 | 106 | 26 | 612 | 148 | 650 | 266 | 221 | 75 | 5776 | 334 | 233 |
| **S2** | 31 | 1294 | 76 | 19 | 139 | 162 | 11 | 554 | 345 | 895 | 5 | 82 | 5598 | 205 | 34 | 29 | 83 | 1530 |
| S3 | 77 | 1073 | 19 | 11 | 47 | 214 | 7 | 54 | 37 | 218 | 8 | 951 | 653 | 458 | 26 | 137 | 702 | 181 |
| **S4** | 25 | 916 | 381 | 230 | 987 | 500 | 12 | 48 | 140 | 195 | 7 | 738 | 776 | 117 | 68 | 226 | 548 | 1610 |
| **S5** | 14 | 106 | 16 | 7 | 29 | 32 | 35 | 22 | 6720 | 619 | 38 | 631 | 5948 | 43 | 24 | 491 | 719 | 237 |
| **S6** | 2217 | 624 | 50 | 18 | 125 | 68 | 12 | 54 | 24 | 285 | 26 | 8142 | 44 | 793 | 75 | 104 | 885 | 217 |
| **S7** | 32 | 1411 | 883 | 570 | 1909 | 75 | 21 | 116 | 152 | 197 | 40 | 4842 | 372 | 182 | 98 | 327 | 749 | 79 |
| **S8** | 70 | 45 | 24 | 14 | 285 | 1794 | 13 | 68 | 2252 | 110 | 7 | 524 | 77 | 2486 | 84 | 737 | 169 | 215 |
| **S9** | 11 | 403 | 10 | 3 | 43 | 23 | 1 | 5679 | 34 | 553 | 8 | 37 | 91 | 43 | 10 | 1218 | 52 | 109 |
| **SLE (pre-pandemic)** | | | | | | | | | | | | | | | | | | |
| **SL1** | 23398 | 4840 | 40 | 76 | 506 | 88 | 68 | 69 | 144 | 281 | 18 | 111 | 1145 | 683 | 31 | 698 | 699 | 142 |
| **SL2** | 4663 | 19781 | 10928 | 21 | 145 | 132 | 17 | 30 | 274 | 293 | 17 | 58 | 200 | 13378 | 1139 | 1826 | 397 | 208 |
| SL3 | 174 | 64 | 29 | 14 | 64 | 129 | 11 | 15 | 17 | 1102 | 30 | 42 | 130 | 134 | 197 | 232 | 55 | 58 |
| **SL4** | 62 | 1876 | 1647 | 1122 | 3379 | 61 | 34 | 134 | 85 | 405 | 17 | 4265 | 882 | 476 | 1541 | 549 | 153 | 7342 |
| **SL5** | 27199 | 10533 | 106 | 42 | 259 | 154 | 37 | 171 | 563 | 20933 | 525 | 667 | 999 | 14812 | 414 | 736 | 2582 | 900 |
| **SL6** | 36 | 1539 | 132 | 89 | 742 | 160 | 5416 | 114 | 332 | 2846 | 72 | 298 | 1412 | 152 | 674 | 216 | 244 | 179 |
